# Predicting the effort required to manually mend auto-segmentations

**DOI:** 10.1101/2024.06.12.24308779

**Authors:** Da He, Jayaram K. Udupa, Yubing Tong, Drew A. Torigian

**Author notes:** <u>Corresponding author:</u> Jayaram K. Udupa PhD, Medical Image Processing Group, 602 Goddard building, 3710 Hamilton Walk, Department of Radiology, University of Pennsylvania, Philadelphia PA 19104.

## Abstract

Auto-segmentation is one of the critical and foundational steps for medical image analysis. The quality of auto-segmentation techniques influences the efficiency of precision radiology and radiation oncology since high-quality auto-segmentations usually require limited manual correction. Segmentation metrics are necessary and important to evaluate auto-segmentation results and guide the development of auto-segmentation techniques. Currently widely applied segmentation metrics usually compare the auto-segmentation with the ground truth in terms of the overlapping area (e.g., Dice Coefficient (DC)) or the distance between boundaries (e.g., Hausdorff Distance (HD)). However, these metrics may not well indicate the manual mending effort required when observing the auto-segmentation results in clinical practice.

In this article, we study different segmentation metrics to explore the appropriate way of evaluating auto-segmentations with clinical demands. The mending time for correcting auto-segmentations by experts is recorded to indicate the required mending effort. Five well-defined metrics, the overlapping area-based metric DC, the segmentation boundary distance-based metric HD, the segmentation boundary length-based metrics surface DC (surDC) and added path length (APL), and a newly proposed hybrid metric Mendability Index (MI) are discussed in the correlation analysis experiment and regression experiment. In addition to these explicitly defined metrics, we also preliminarily explore the feasibility of using deep learning models to predict the mending effort, which takes segmentation masks and the original images as the input.

Experiments are conducted using datasets of 7 objects from three different institutions, which contain the original computed tomography (CT) images, the ground truth segmentations, the auto-segmentations, the corrected segmentations, and the recorded mending time. According to the correlation analysis and regression experiments for the five well-defined metrics, the variety of MI shows the best performance to indicate the mending effort for sparse objects, while the variety of HD works best when assessing the mending effort for non-sparse objects. Moreover, the deep learning models could well predict efforts required to mend auto-segmentations, even without the need of ground truth segmentations, demonstrating the potential of a novel and easy way to evaluate and boost auto-segmentation techniques.

## 1. Introduction

### 1.1 Background

Image segmentation is performed to delineate target objects in an image of interest [1], which has been a topic of main interest in the general image processing field since the 1960s [2, 3]. In the domain of medical images, tomographic 3D image segmentation has flourished with the advent of computed tomography (CT) after the 1970s [4, 5]. Medical image segmentation plays a particularly important role in dozens of medical image analysis applications, such as 3D reconstruction [6], quantitative measurement [7], and radiotherapy planning [8]. Many early segmentation approaches were purely image-based, which often required human interaction for the method itself or considerable post hoc correction. To improve time-efficiency of segmentation, auto-segmentation techniques emerged. Among these, prior-knowledge-based approaches consisting of object-model-based methods [9, 10, 26] and atlas-based methods [11, 12] have been widely studied. Recently, deep learning-based prior knowledge methods [13-16] have taken off to significantly improve the accuracy and efficiency of medical image segmentation over past methods for use in numerous practical applications [17-19, 33].

Meaningful metrics to evaluate auto-segmentation methods are critical for developing auto-segmentation algorithms that are clinically effective and useful. An ideal metric should effectively express: *P1. Accuracy*: the accuracy of the segmentation relative to a given reference ground truth; *P2. Acceptability*: the clinical acceptability of the segmentation for the application at hand; *P3. Computational efficiency*: the computational time cost of the segmentation method (typically, this involves the cost of model building/training and the per-study cost of performing segmentation); *P4. Mending efficiency*: the post hoc human time required to correct the auto-segmentation of the method. Poor accuracy, inadequate clinical acceptability, low computing efficiency, and low post hoc correction efficiency can affect the quality and utility of an auto-segmentation algorithm in clinical practice. Typically, manual inspection and correction is usually required following any auto-segmentation utilized clinically where clinical experts mend any unacceptable auto-segmentation errors (including both false positive and false negative errors) in a slice-by-slice manner. Among these properties, P1, P2, and P4 behave differently than P3 in that they need a human expert involvement to provide the reference/judgement while P3 can be established totally objectively independently of any reference expert<u>^1^.</u> The challenges in segmentation evaluation all stem from P1, P2, and P4 since they all have a subjective component and require the expensive time of experts. Due to these reasons, P3 will not be our concern in this paper. Since P4 requires an expert to closely scrutinize and correct the auto-segmentation, we argue that it subsumes P1 and P2. This is due to the fact that we expect the expert to perform mending to agree with the ground truth they have in mind, and resultantly, the mended segmentation to be highly acceptable to them. In other words, P4 is the most crucial and practically useful characteristic of a segmentation metric.

Following the above argument, we seek answers to the following questions in this paper: (Q1) How effective are most commonly-used metrics like the Dice Coefficient (DC) [20] and Hausdorff Distance (HD) [21] in portraying/predicting mending efficiency? (Q2) Can metrics be explicitly defined to portray/predict mending efficiency accurately? (Q3) Can metrics be implicitly defined through deep learning models to portray/predict mending efficiency accurately? As illustrated in Fig. 1, although both auto-segmentations in (a) obtain the same DC of 0.80, the errors are different in terms of mending efforts required for correction. Almost all of the false positive errors in the first example of (a) are closely attached to most segments of the true positive region, while error regions in the second example are roughly clustered in two regions that can be easily distinguished from the true positive region. A clinical expert would likely need to spend a lot more time in correcting the first example than the 2^nd^. On the other hand, both examples in pair (b) obtain the same HD of 82 pixels. Yet, it is likely that the expert will require more time to mend the second auto-segmentation result compared to the first one that requires only a single mouse operation for removing the isolated false positive error region. Explicitly formulated closed-form metrics such as DC and HD are designed to indicate deviation of auto-segmentations from ground truth segmentations. Mending effort on the other hand depends on user behavior, variation of this behavior among institutions and experts, correction software tools used, etc., which are hard to capture via simple quantification of the overlap or distance between segmentation masks.

**Figure 1.**
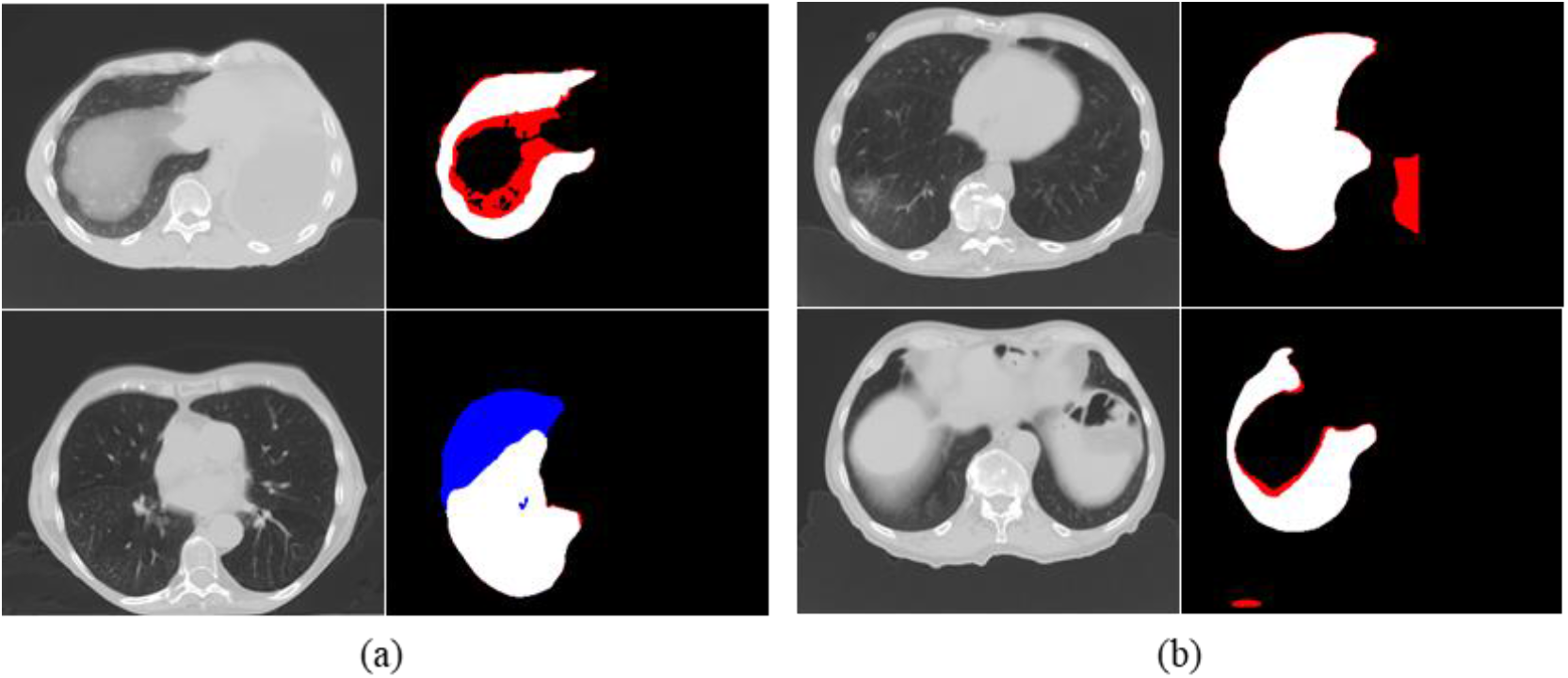
Some thoracic CT slices with their auto-segmentation masks. White = true positive region; red = false positive region; blue= false negative region. Both auto-segmentations in (a) have a DC= 0 .80, while both auto-segmentations in (b) obtain a HD = 82 pixels.

From the clinical workflow perspective, it would be very useful if we could predict the mending effort required for any given study accurately. For example, in a Radiation Therapy (RT) unit, this would allow appropriately managing workflow by identifying cases that require considerable mending effort vs. those that require minimal effort and accordingly distributing pending mending tasks among dosimetrists and oncologists. It is, therefore, highly desirable to design a “metric” either as an explicit formulation or via implicit modeling to be able to predict the mending effort required for a given study just from the patient study and the associated auto-segmentation.

### 1.2 Related work

Existing segmentation metrics can be grouped into three main types. The first type is based on areas (or volumes for 3D segmentations), which considers the areas of both auto-segmentations and GT segmentations to obtain a ratio of the true positive (TP) auto-segmentation areas. Area-based metrics, such as DC [20] and Jaccard Index (i.e., Intersection over Union) [27], have been widely used in numerous medical image segmentation studies. By considering attention to different errors, [28] mentioned that different types of area errors should be weighted differently in different applications. However, area-based metrics do not consider the shape or the position of errors, which will influence mending behaviors (e.g., Fig. 1(a)).

The second type is based on boundary distance-based metrics, including HD [21], average symmetric surface distance, and root mean square symmetric surface distance [29, 30], which are commonly used. These metrics are calculated by comparing the segmented mask boundaries of the auto-segmentation and the GT and defining a distance between the two irregular sets. Only pixels on the boundary contribute to the metric value, while the number of pixels inside the boundary is negligible for calculating the metric value. As a result, these metrics cannot well characterize spike-shape errors and isolated errors (e.g., Fig. 1(b)), which can be easily corrected with segmentation annotation software.

The third type is based on boundary length-based metrics. Different from boundary distance-based metrics (e.g., HD) that focus on the distance between pixel pairs from two boundaries, boundary length-based metrics are calculated by counting the pixels along the boundaries. All pixels along the boundaries can be classified as TP, false positive (FP), true negative (TN), or false negative (FN) elements, and thus some ratios can be calculated according to the quantities of different types of elements, such as surDC [22] and APL [23]. Boundary length-based metrics might better approach the correction behaviors than the other two types of metrics since human experts draw correction pixels along the boundaries. However, there are also some drawbacks of these metrics. For example, the isolated error regions as shown in Fig. 1(b) cannot be well distinguished from the attached error regions using these metrics, thereby influencing the potential to estimate the mending effort for auto-segmentations.

There are very limited studies that try to figure out the relationship between segmentation metrics and clinical value. Based on a reader study, [31] recorded a qualitative acceptability score (AS) for segmentations and estimated metric-AS relationships. A linearization concept was proposed to improve the uniformity of the relationships. Although AS can reflect the clinical value to some extent, a more quantitative approach is desired to express the clinical value. In contrast, [23] calculated the correlations between time-saving and various segmentation metrics. Time-saving is a proper index for measuring the relationship between segmentation metrics and clinical value. However, there are some limitations in [23]. On the one hand, the segmentation metrics compared in [23] can be further improved to better simulate subjective correction, thereby better indicating mending time-saving. On the other hand, a more detailed analysis is expected, including the multi-center study, the study for multi-expert labeling, and the discussion about different behaviors for different objects.

However, it seems that works on how to precisely predict or estimate the mending effort are deficient. [23] shows some correlations based on segmentation metrics, which can be roughly regarded as linear predictors, but detailed analyses and further explorations are needed. Although it is important to determine which segmentation metric could robustly indicate the mending effort, it is also important to explore ways to accurately estimate the effort required for mending a given auto-segmentation. To the best of our knowledge, previous research has been lacking in terms of providing ways to predict the mending effort without auto-segmentation GT, which may be very helpful in various scenarios involving auto-segmentation.

In this paper, we discuss the above issues and propose methods to answer these questions to some extent. Four existing metrics, DC, HD, Surface DC (surDC) [22], and Added Path Length (APL) [23] are considered for this purpose. In addition, a new metric, Mendability Index (MI), is proposed in this work. Both correlation analysis and regression analysis are applied to these five metrics to study their relevance to mending effort. More interestingly, besides these explicitly defined metrics, three deep learning pipelines for predicting the mending effort are explored as implicit metrics, providing more accurate and flexible solutions in an implicit manner. Therefore, this paper introduces the following contributions. Firstly, detailed analyses between existing segmentation metrics with mending effort are conducted based on multi-center data for seven objects. Second, as a supplement to existing metrics, the concept of MI and its variant are proposed. Third, deep learning-based methods are verified to be feasible for mending effort estimation, even without the requirement for GT.

Some preliminary results of our study have been presented in a conference report [32]. Compared with [32], this current article includes significant improvements in several respects. Firstly, the newly proposed metric MI with the explicit formula has been further developed by combining the segmentation boundary distance term. Second, the analysis for metrics has been enriched, including more existing metrics, more objects, and regression analysis. Moreover, deep learning-based regression analysis has been introduced in this article, demonstrating the potential of implicit segmentation metrics (i.e., metrics implicitly defined by millions of parameters) for indicating the mending effort.

### 1.3 Outline of approach

This article tries to better discuss segmentation metrics for estimating mending effort in detail, and is organized as shown in Fig. 2. In Section 2, we introduce the collected dataset for evaluation and all of the compared metrics. Specifically, we propose two groups of predictive models: explicit and implicit models. Explicit models make use of analytical metrics, including DC, HD, surDC, APL, and a MI-based new metric. We build a model for each metric to predict mending effort required by making use of given segmentation GT, auto-segmentation, and effort data. For implicit models, we propose 3 deep learning-based models trained on: (i) GT and auto-segmentation, (ii) GT, auto-segmentation, and CT image, (iii) auto-segmentation and CT. In Section 3, we analyze the predictive accuracy of all models based on multi-institutional clinical data and the actual mending time needed by experts to correct auto-segmentations. The observations of both correlation analysis and classical regression analysis can be divided for sparse and non-sparse objects. Moreover, the results of the deep learning-based regression are also elaborated, showing better performance than metric-based classical regression. In Section 4, we discuss the significance and limitations of this study. In Section 5, the findings from the experiments are summarized.

**Fig. 2.**
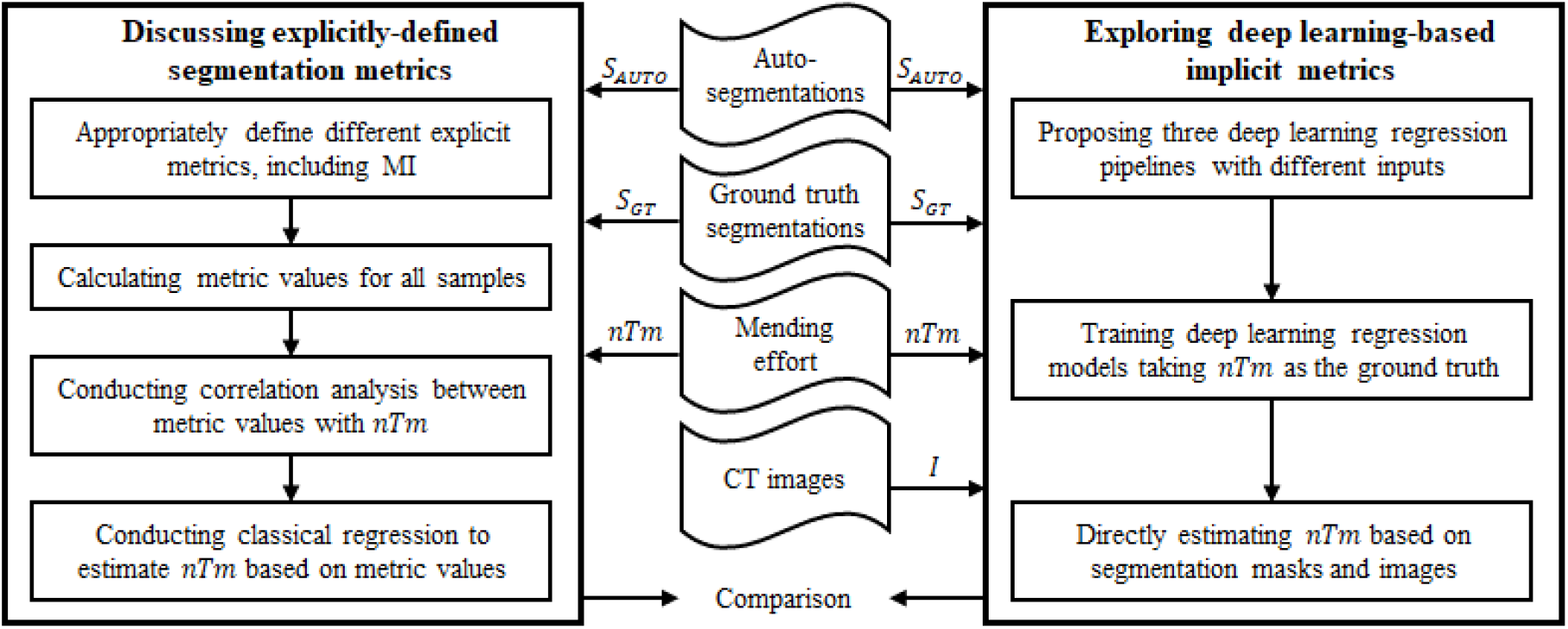
A schematic representation of the segmentation metric discussion.

## 2. Method

### 2.1 Data sets

To study the relationship between different metrics with the effort required for editing auto-segmentations, a dataset was collected from 3 institutions [33]. Adult subjects with known or suspected cervical or thoracic malignancy undergoing clinical CT imaging for radiation therapy planning are included in this dataset. For generalizability, various parameters for subject demographics, subject condition, scanner models, and imaging settings were considered during the data collection. After acquiring the CT images, GT segmentations were independently annotated following standardized definition of objects of interest [33]. Clinical contours and manually edited auto-segmentation contours were created by different dosimetrists from the 3 institutions. Details of the image data, the GT segmentation, and the data on mending effort are as follows.

#### Image data

To demonstrate the performance of different segmentation metrics, this work includes 7 objects: left submandibular gland (LSmG), mandible (Mnd), cervical spinal cord (CtSC), orohypopharyngeal constrictor muscles (OHPh), cervical trachea (CtTr), right lung (RLg), and heart (Hrt). RLg and Hrt can be classified as non-sparse objects, which have clear boundaries and compact shapes, and are usually easier to segment. In contrast, the other five objects are sparse objects, showing more variable, less compact, and often smaller shapes, usually leading to poor segmentations. There are other differences between sparse and non-sparse objects. For example, clinical experts may show higher subjective acceptability for auto-segmentations for sparse objects than for non-sparse objects with the same DC [33]. In addition, different neural network architectures should be considered when designing the deep learning-based auto-segmentation methods. Therefore, it is very important to contain both types of objects in this general study. The numbers of collected subjects for each object are presented in Table 1.

**Table 1.**
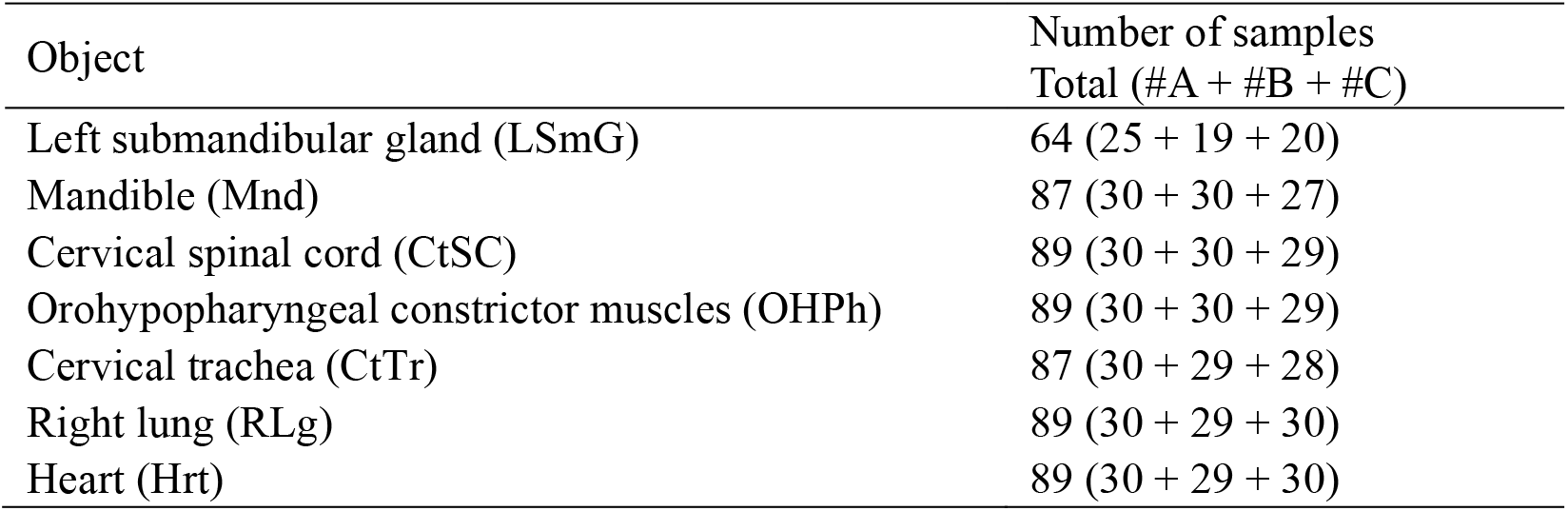
Number of collected segmentation samples for each object. #A, #B, and #C indicate the numbers of samples from each institution.

#### GT segmentation

After obtaining the CT images for both the thorax region and the neck region, the GT segmentations were annotated by our group (i.e., instead of the three clinical institutions) following the precise and standardized definition of objects, which is (i) anatomically relevant for the application and valid in subjects; (ii) feasible to implement; and (iii) able to obtain high recognition accuracy [33]. The same object definitions were applied when building the auto-segmentation methods used in this study. Therefore, values of various segmentation metrics can be calculated based on the GT and auto-segmentations.

#### Data on mending effort

The CT images were handled by a hybrid intelligence system [33] to generate auto-segmentation results. These auto-segmentation results were sent back to the three institutions for manual correction following their routine clinical practice. Note that the dosimetrists/radiation oncologists in different institutions did not work on the same scans, although they corrected the same 7 objects. Scans with the auto-segmentations were distributed to these institutions according to their original sites. The time cost for mending each sample at the corresponding institution was recorded. Therefore, for each sample of each object, the effort required for manually mending was quantified by the mending time (*Tm*). It is possible to evaluate the relationship between segmentation metric values with *Tm*.

However, *Tm* labeling can be evidently influenced by institution standards, labeling tools, and personal experience. In other words, even with the same CT image and auto-segmentation, different readers may label different *Tm* values. Therefore, *Tm* cannot reflect the general quality of auto-segmentations from this multi-center data. As shown in Table 2, the recorded maximum *Tm*, mean *Tm*, and standard deviation (std) *Tm* values vary a lot among different experts from different institutions.

**Table 2.** Human readers’ behaviors in terms of their *Tm* labeling for all objects.

| Institution | Reader alias | Minimal $Tm$ (min) | Maximal $Tm$ (min) | Mean (std) $Tm$ (min) |
| --- | --- | --- | --- | --- |
| Institution A | Expert I | 0.0 | 4.00 | 0.44 (0.77) |
|  | Expert II | 0.0 | 7.00 | 0.90 (1.18) |
| Institution B | Expert III | 0.0 | 3.87 | 0.89 (0.68) |
| Institution C | Expert IV | 0.05 | 11.23 | 2.09 (1.69) |
|  | Expert V | 0.0 | 12.02 | 1.33 (2.46) |

In this regard, for the editing time recorded by each expert, we normalized *Tm* values according to the maximal *Tm* value recorded by this expert. The normalized *Tm* (*nTm*) can be expressed as follows:

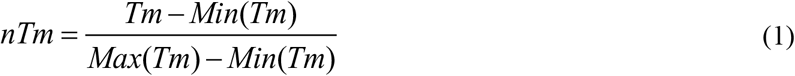

Note that we took *Min*(*Tm*) as 0 for all experts since there can be no modifications on the good enough auto-segmentation result and since it is convenient to predict the actual *Tm* by multiplying by *Max*(*Tm*) in practice. Therefore, *nTm* could well reflect the mending effort of each individual among different objects. The following experiments are conducted by studying the relationship between segmentation metrics with *nTm*.

### 2.2 Analytic metrics

Common metrics often evaluate the deviation of auto-segmentation from GT by treating them as 3D entities. However, it is important to point out that human readers observe and correct auto-segmentations in a slice-by-slice manner. In other words, although the auto-segmentation for an object is a 3D volume with many 2D slices and the *nTm* label was labeled according to the whole 3D volume, clinical experts actually did corrections for each 2D slice and accumulated the mending time. It is not practical to directly modify a 3D shape in clinical practice. As a result, when applying existing metrics for estimating the mending effort, considerations of the above human manner are needed for a fair comparison. As such, we used 2D versions integrated over all slices to create metrics that match with human behavior.

#### Dice Coefficient

DC has been widely used in massive segmentation studies, which is usually defined in 3D space for medical segmentation as follows:

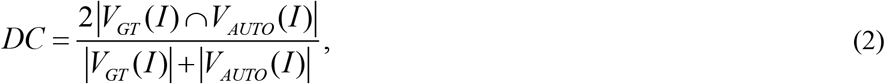

where *I* indicates the 3D CT image consisting of multiple slices *Is, V*_*GT*_ indicates the GT segmentation, *V*_*AUTO*_ refers to the auto-segmentation, and |·| represents the cardinality of the set.

However, the 3D version of DC mixes all of the voxels from all slices to obtain the metric score, and does not consider the manual annotation procedures. Therefore, we apply mean DC (i.e., mDC) in this study to describe the 3D segmentation using the concept of DC, which is defined as follows:

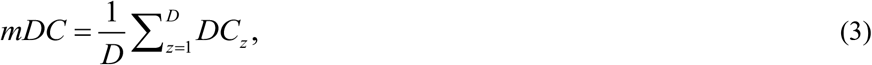

where *D* indicates the number of valid slices, and *DC*_*z*_ refers to the 2D DC of the *z*^*th*^ slice *Is*.

#### Hausdorff Distance

Different from DC, HD describes the distance between segmentation boundaries. A larger HD is expected to represent poorer auto-segmentation quality. In the 2D plane, if *B*_*GT*_*(Is)* corresponds to the boundary of GT segmentation *S*_*GT*_*(Is)* and if *B*_*AUTO*_*(Is)* corresponds to the boundary of auto-segmentation *S*_*AUTO*_*(Is)*, HD can be defined using:

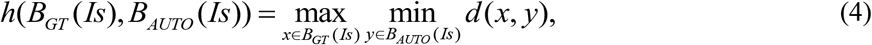

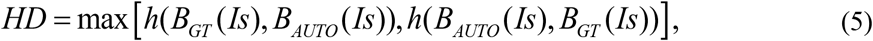

where *d(x, y)* indicates the Euclidean distance between two pixels *x* and *y*.

HD is not a normalized score, and can be as large as the maximal distance between any two pixels along *B*_*GT*_*(Is) and B*_*AUTO*_*(Is)*. Therefore, by considering the slice-by-slice mending behavior, we utilize the sum of 2D HD values (i.e., sHD) to evaluate the mending effort for the whole object. sHD can be expressed as follows:

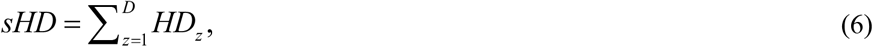

where *HD*_*z*_ refers to the 2D HD of the *z*^*th*^ slice.

#### Surface Dice Coefficient

Compared with area-based metrics (e.g., DC) and boundary distance-based metrics (e.g., HD), boundary length-based metrics seem more appropriate when simulating the manual correction behavior. This is because human experts usually draw new boundaries or erase existing boundaries, instead of directly modifying the area or adjusting the boundary distance when mending the auto-segmentations. As a result, boundary length-based metrics need to be discussed in this paper.

As shown in Fig. 3, pixels along *B*_*GT*_*(Is)* and *B*_*AUTO*_*(Is)* can be classified into 3 types. Pixels that are both on *B*_*GT*_*(Is)* and *B*_*AUTO*_*(Is)* belong to TP predictions (e.g., as indicated by the green triangles in Fig. 3(a)). Similarly, there are also two types of wrong predictions, FP and FN pixels, corresponding to the predicted boundary outside of *S*_*GT*_*(I)* and inside of *S*_*GT*_*(I)*.

**Fig. 3.**
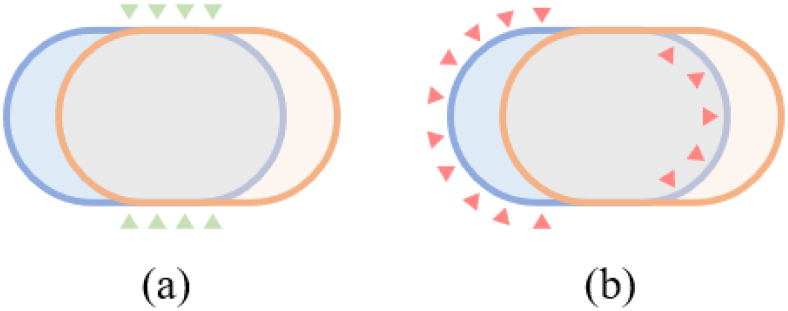
Illustrations of surDC and APL metrics. The blue region and the orange region indicate the ground truth segmentation and the auto-segmentation, respectively. The green triangles in (a) refer to the TP segments. The red triangles in (b) refer to the FP and FN segments along the ground truth boundary.

surDC [22] is a segmentation metric that focuses on the proportion of TP boundary segments in pixels (i.e., the overlapped part that does not need further correction). surDC can be defined as follows:

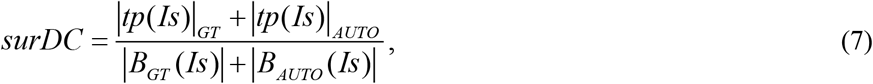

where |*tp*(*Is*)|_*GT*_ corresponds to the number of TP boundary pixels along *B*_*GT*_*(Is)*, and |*tp*(*Is*)|_*AUTO*_ corresponds to the number of TP boundary pixels along *B*_*AUTO*_*(Is)*.

As there could be an acceptance tolerance between *B*_*GT*_*(Is)* and *B*_*AUTO*_*(Is)* (i.e., some trivial errors could be ignored in practice),|*tp*(*Is*)|_*GT*_ may be not strictly equivalent to |*tp*(*Is*)|_*AUTO*_. In addition, surDC can be a metric for the whole 3D segmentation, since all the length terms in Eq. (7) can be accumulated slice-by-slice, which is reasonable considering the auto-segmentation mending behavior.

#### Added Path Length

Although APL [23] is also a boundary length-based metric, it is different from surDC. The latter pays attention to only the correctly predicted boundaries that do not need further mending, while the former focuses on the wrongly predicted boundaries. As shown in Fig. 3(b), APL counts the number of FP and FN boundary pixels along *B*_*GT*_*(Is)*, which can be expressed as:

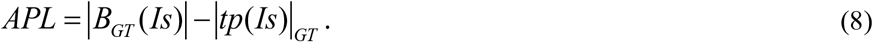

Similar to HD, since APL is not a normalized metric, we can add up slice-wise APL values to obtain sAPL for estimating the mending effort for the whole object, as shown in Eq. (9):

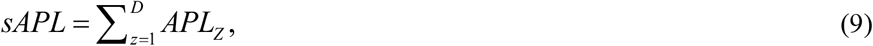

where *APL*_*z*_ refers to the APL of the *z*^*th*^ slice *Is*.

#### Mendability Index: Types of segmentation error

Human readers observe auto-segmentations subjectively and correct errors when finding unacceptable auto-segmentation pieces. With the help of annotation software tools, those FP and FN errors are mended until the segmentation becomes acceptable. Specifically, readers may have different behaviors when faced with different errors, which can be classified into three types.

Let the region shown in blue in Fig. 4(a) represent *S*_*GT*_*(Is)* and the region shown in orange represent *S*_*AUTO*_*(Is)*. Note that the blue mask is slightly hidden by the orange mask. Let *S*_*0*_*(Is)* be the TP region of *S*_*AUTO*_*(Is)*. Consider the 3 FP segments of *S*_*AUTO*_*(Is)* denoted *S*_*1*_*(Is), S*_*2*_*(Is)*, and *S*_*3*_*(Is)* and the 3 FN segments *S*_*4*_*(Is), S*_*5*_*(Is)*, and *S*_*6*_*(Is)*.

**Fig. 4.**
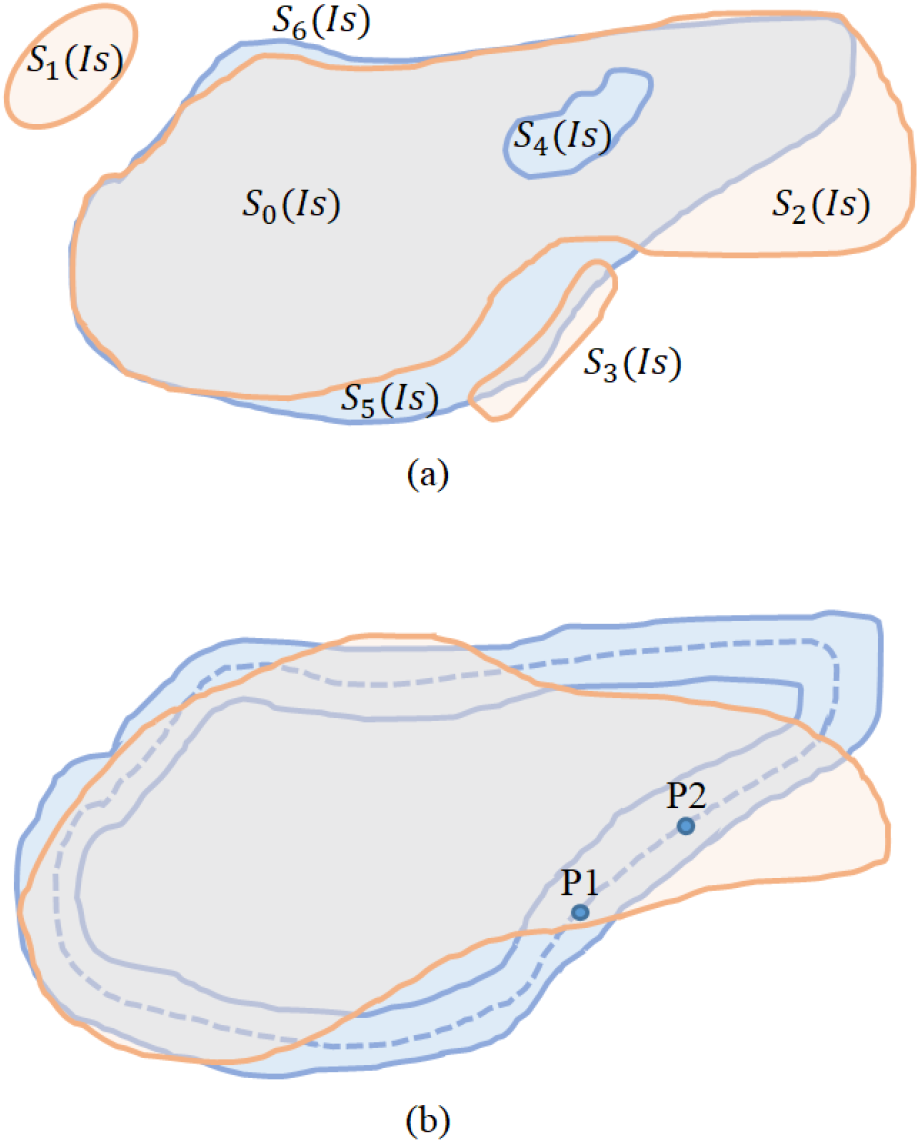
Illustrations of different segmentation components. The blue region and the orange region indicate the GT segmentation and the auto-segmentation, respectively. (a) Examples of TP component (*S*_*0*_), three types of FP errors (*S*_*1*_*∼*S_3_), and three types of FN errors (*S*_*4*_*∼*S_6_). (b) The strict ground truth segmentation boundary (the dashed blue line), the tolerated GT segmentation boundary (the solid blue lines), and the auto-segmentation boundary (the solid orange line).

We identify three types of FP components that differ in cost for mending. (T1) Segments of *S*_*AUTO*_*(Is)* like *S*_*1*_*(Is)* which are not connected to *S*_*GT*_*(Is)* cost very little for mending since they can be removed by a simple action like a mouse click. (T2) Segments like *S*_*2*_*(Is)* which are close and connected to *S*_*GT*_*(Is)*, but otherwise are thick and not subtle incur more time for careful mending than T1-type components, the cost depending on the length of the boundary segment of *S*_*GT*_*(Is)* that is adjacent to the FP component. (T3) Segments like *S*_*3*_*(Is)* take the longest time to mend which are very subtle and close to the boundary of *S*_*GT*_*(Is)*. As for FNs, we can identify three types of components for FNs. (T1) Components like *S*_*4*_*(Is)* that can be easily included. (T2) Components like *S*_*5*_*(Is)* which take an intermediate amount of effort to fix. (T3) Segments like *S*_*6*_*(Is)* which lie close to the boundary of *S*_*GT*_*(Is)* and hence take more time to fix carefully.

#### Mendability Index: Definition

In practice, the correction for T2 and T3 errors is usually segmentation boundary-based, which might coincide with the motivation behind boundary length-based metrics. Therefore, it is reasonable to evaluate T2 and T3 errors by considering different segments along *B*_*GT*_*(Is)*. Specifically, given an FP sub-region *S*_*i*_*(Is)* (e.g., *S*_*2*_*(Is)* in Fig. 4(a)), the segment of *B*_*GT*_*(Is)* to which *S*_*i*_*(Is)* is attached is called an FP boundary segment and expressed as *fpi(Is)*. Similarly, we can also define a FN boundary segment *fni(Is)* (e.g., for *S*_*5*_*(Is)*). Then, the proportions of *fpi(Is)* and *fni(Is)* can be used to evaluate the T2 and T3 errors. In addition, considering the different correction behavior for T1 errors, the number of T1 sub-regions can also contribute to the mending effort. In this way, the MI for auto-segmenting an object *O* in image *I* can be defined as:

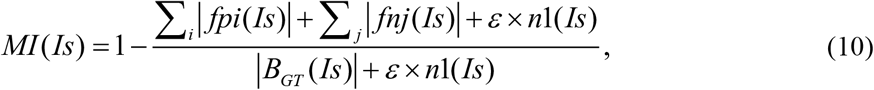

where |· | indicates the length of the boundary segment, *n1(Is)* denotes the number of T1 sub-regions, and *ε* is the positive coefficient of *n1(Is)*. A higher MI value may indicate better auto-segmentation.

Different from many existing metrics, to improve the completeness of the metric definition, the boundary pixel edges instead of boundary pixels are applied to calculate |·| terms in Eq. (10). The latter definition (i.e., boundary pixel-based) may encounter some topological issues. For example, the “Jordan property” of the digital boundary describes that the boundary should be closed so that it divides image space into two disjoint parts, such that any path taken from the interior to the exterior intersects the boundary. However, in the case of identifying boundaries by pixels, defining closed or Jordan boundaries becomes very challenging and impossible in degenerate cases of thin and subtle objects [24, 25, 26]. As shown by the examples in Fig. 5(b), when the segmentation mask is thin, small, and subtle, we cannot well define the segmentation boundary using pixels. In contrast, a sequence of pixel edges indicated by the dark blue dashed line makes sense, even for a single pixel.

**Fig. 5.**
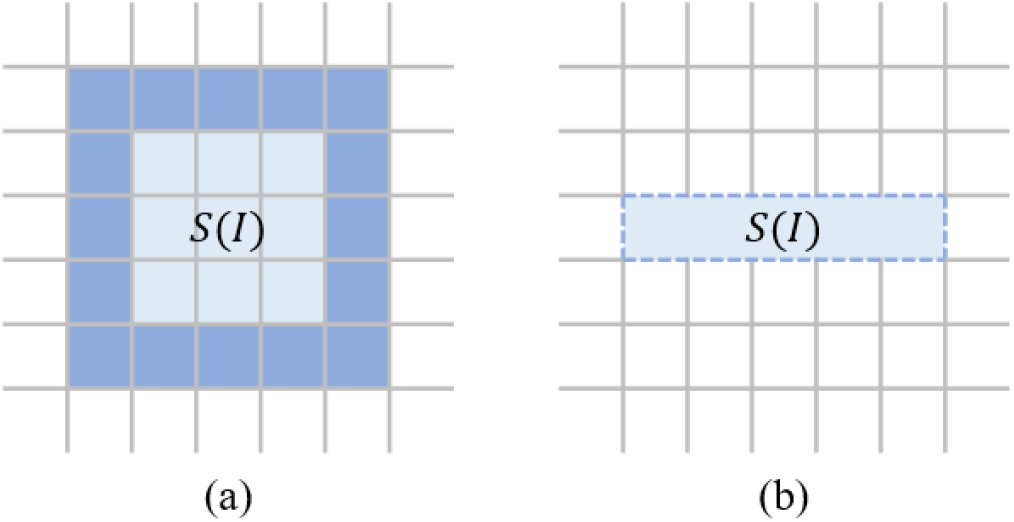
Illustrations of different boundary definitions. Each cell indicates an image pixel. Colored cells (light and dark blue) correspond to the segmentation mask. Dark blue shows the segmentation boundary. (a) The boundary is a set of pixels. (b) The boundary is a set of pixel edges indicated by the dark blue dashed line.

#### Tolerance parameter to decide acceptable *B*_*GT*_*(I)*

When labeling or correcting the segmentation boundaries, human variations and boundary uncertainties can influence the precise results. The former issue comes from subjective observations, while the latter is related to finite resolution, image blur, noise, similar image intensity properties with adjacent tissues, etc. Therefore, when comparing *B*_*AUTO*_*(Is)* with *B*_*GT*_*(Is)*, if the gap between the two segments from *B*_*AUTO*_*(Is)* with *B*_*GT*_*(Is)* is small enough, manual correction is not necessary and the auto-segmentation segment is acceptable. To simulate the tolerated and fuzzy subjective behavior for deciding the acceptable boundary, as shown in Fig. 4(b), a tolerance parameter *δ* is applied to decide the acceptable *B*_*GT*_*(Is)*. For example, in Fig. 4(b), in addition to the strict boundary of *S*_*GT*_*(Is)* (i.e., the dashed blue line), there are two solid blue lines showing the tolerance range, which have the distance of *δ* to the dashed blue line. Each of the elements (i.e., pixel edge) along the strict *B*_*GT*_*(Is)* can obtain the TP status as long as there are any auto-segmentation boundary elements within the distance of *δ*. Note that the distance between two pixel edges is defined by the distance between the corresponding edge midpoints. For each of the pixel edges along the strict *B*_*GT*_*(Is)*, a loop search can be implemented to check if any nearby pixel edge belongs to *B*_*AUTO*_*(Is)*. As a result, the representative pixel edge P1 corresponds to an acceptable result, while the representative pixel edge P2 contributes to |*fpi*(*Is*)|.

In this way, the fluctuations of human behaviors and the uncertainties of the imaging for manually delineating organs have been considered, resulting in a more practical MI metric. Although the classification of *fpi(Is)* and *fni(Is)* has been improved by introducing a tolerance, *fpi(Is)* and *fni(Is)* are still sub-sets of *B*_*GT*_*(Is)* and Eq. (10) still holds.

#### Mendability Index: Implementation for a 3D volume

The terms in Eq. (10) are defined in 2D slices because human readers usually observe and correct the 3D auto-segmentations in a 2D slice-by-slice manner. However, Eq. (10) can also be applied to evaluate the whole 3D volume by accumulating |*B*_*GT*_ (*Is*)|, |*fpi*(*Is*)|, |*fni*(*Is*)|, and *n1(Is)* for all slices before calculating the final MI.

#### Mendability Index: Modulation with Hausdorff Distance

Although the correction operation is consistent with the segmentation boundary length-based metric concept, our preliminary experiments show that segmentation boundary distance may also be related to the mending effort. Therefore, inspired by previous studies that fuse different metrics [29, 34-36], we combine MI and HD with the following definition:

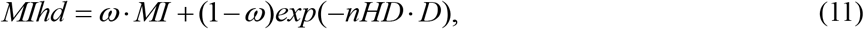

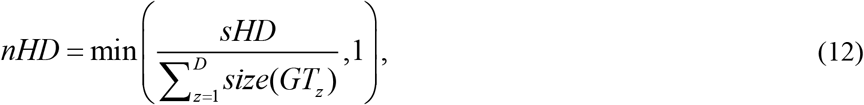

where *size(GT*_*z*_) indicates the diameter of the minimal enclosing circle of *S*_*GT*_*(I)* and *ω* is a coefficient to balance the two terms.

The improved version, MIhd, still has the value range of 0∼1 with a positive correlation with the auto-segmentation quality. There are two motivations behind Eq. (11). Similar to the idea of the tolerance parameter, the boundary distance may influence the general observation. Yet, as different subjects may have different organ sizes, instead of directly applying sHD, the normalized version nHD and the number of valid slices *D* are introduced to better consider individual differences. Therefore, MIhd may have the potential to describe more complicated variations and will be applied in the following experiments.

### 2.3 Explicit models

With different motivations, the above analytic metrics mDC, sHD, surDC, sAPL, and MIhd can be used to evaluate the auto-segmentation results by comparing them with the GT segmentations. To study their performance for indicating the mending effort in practice, we assess their correlations with the mending effort (i.e., *nTm*) and build explicit models using analytic metrics to estimate *nTm* values.

#### Correlation analysis

Since we have both GT segmentation and auto-segmentation for each sample in terms of each object, we are able to calculate the corresponding analytic metric values. For example, when studying the object Hrt, 89 mDC values can be obtained. Together with the recorded 89 *nTm* values for Hrt, the correlation between these two sets can be calculated, which is also feasible for sHD, surDC, sAPL, and MIhd. By comparing the correlation coefficients from different analytic metrics for the same object, we can study the consistency between manual mending effort with the well-defined analytic metrics.

#### Predictive models

In addition to the correlation analysis showing the linear trend, it would be better to check exactly how well these analytic metrics can estimate the clinical mending effort. Therefore, predictive models were built for each analytic metric for each object. Taking Hrt as the example, the 89 pairs of mDC values and *nTm* values were divided into a training set and a testing set. Predictive models with the mDC value as the input and the *nTm* value as the target were trained on the training set and evaluated using the testing set. Beyond this example, we built predictive models for each of the five analytic metrics for each of the seven objects. For fair comparison with limited data, the training-testing set splitting follows a five-fold cross-validation manner. Two classical regression methods, linear regression model and support vector regression (SVR) model [39, 40], were selected based on some preliminary experiments to build the explicitly predictive models based on analytic metrics.

### 2.4 Implicit models based on deep learning

#### Motivation

As introduced above, different types of segmentation metrics are built based on explicitly well-defined formulas to focus on different expressions of errors, which may result in different performances when estimating the mending effort. However, it is not easy to perfectly consider all correction behaviors and to adaptively fit different objects and different human experts when designing such formulas.

In recent years, we have witnessed the booming development of deep learning for numerous applications. In addition to the auto-segmentation algorithms, image recognition, image-based parameter regression, and image-based measurement tasks are also widely studied, showing the great potential of implicitly fitting using neural networks. Therefore, we hope to conduct a preliminary study in this work to explore if it is possible to estimate the auto-segmentation mending effort using deep learning methods without explicit metrics.

#### Models

Specifically, based on the collected dataset, three types of models are explored, which are illustrated in Fig. 6.

**Fig. 6.**
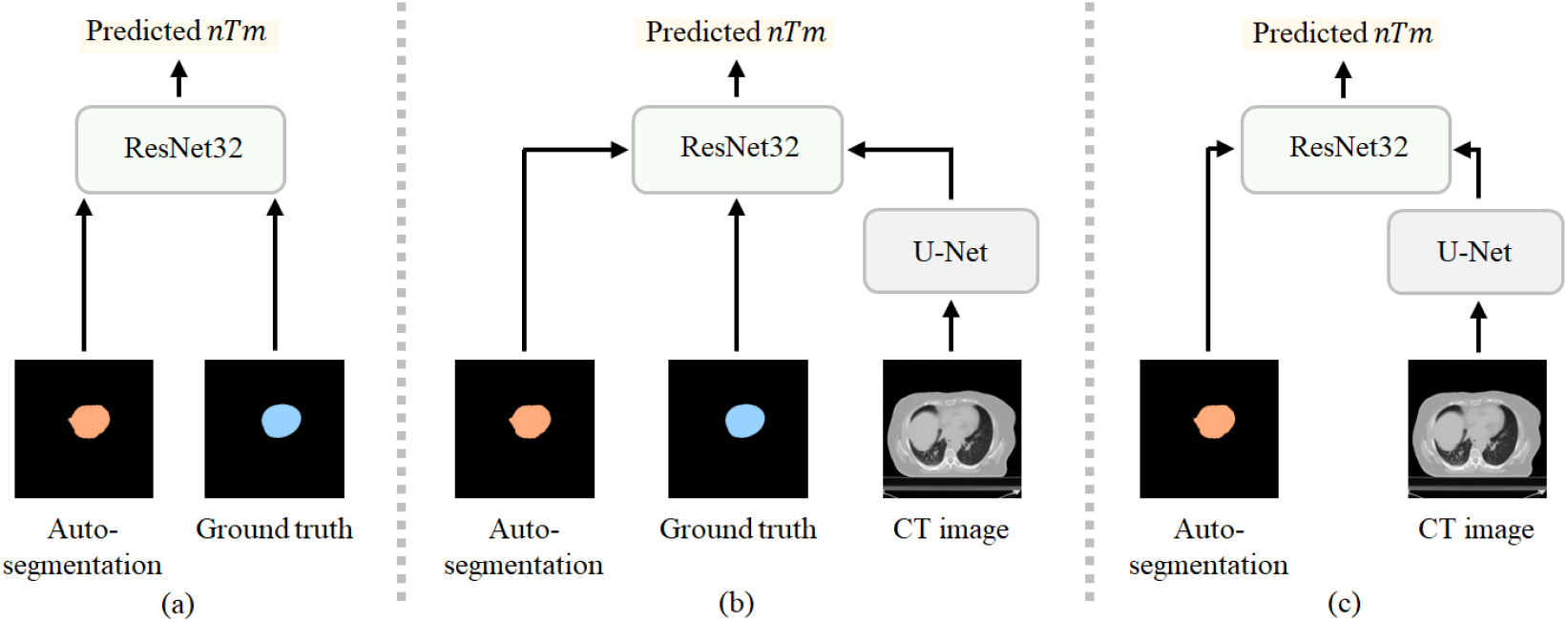
Diagram of deep learning-based regression models. (a) “Auto+GT” model. (b) “Auto+GT+CT” model. (c) “Auto+CT” model. Both the U-Net and ResNet32 architectures have been modified to accommodate the input and output dimensions.

The first deep learning-based model (i.e., Fig. 6(a)) is inspired by the existing well-defined metrics, which calculate the values by comparing the auto-segmentation with the ground truth segmentation. Instead of explicitly defining the formula, the first model (denoted as “Auto+GT” model) applies a popular ResNet32 architecture [37] to adaptatively explore the relationship between the auto-segmentation and the ground truth segmentation. These two types of object segmentation mask arrays are concatenated along the channel dimension as the input of the ResNet32 model. The first layer of the ResNet32 architecture is modified to accommodate the input dimension, and the output layer is also modified for the single-value regression.

Since the convolutional neural network has excellent power for handling complicated information, we also try to input all the information we have to benefit the mending effort estimation. Therefore, in addition to the auto-segmentation mask and the ground truth segmentation mask in “Auto+GT” model, the original CT image is also utilized in the second model (denoted as “Auto+GT+CT” model). As shown in Fig. 6(b), the original CT image is first handled by a U-Net architecture [38] for feature extraction. Then, the extracted features are concatenated with the auto-segmentation and the ground truth segmentation for the final regression using ResNet32. Similarly, the first layer and the last layer of the U-Net and ResNet32 architectures are modified to accommodate the channel dimensions.

We additionally explore the “subtraction” idea in the third model that accepts three types of input information. As shown in Fig. 6(c), the third model (denoted as “Auto+CT” model) removes the branch of ground truth segmentation from the “Auto+GT+CT” model. The first layer and the last layer of U-Net and ResNet32 are modified accordingly to fit the channel dimension. There is a particularly significant motivation behind the “Auto+CT” model. With the “subtraction,” no GT segmentation information is needed. In other words, if this model works, then the manually labeled GT segmentation becomes unnecessary for measuring the auto-segmentations in terms of the required mending effort.

The sigmoid function is applied as the output activation for the three deep learning-based pipelines. Meanwhile, all the models are trained in a supervised manner. The regression ground truth for the supervised learning is the *nTm* value, enabling the network to estimate the effort required for mending the auto-segmentation.

#### Loss function

To train the above deep learning models in a supervised manner, two loss terms are combined in the total loss function. First, the typical L1 loss (ℒ_*L*1_) is utilized for efficient convergence. However, we observe that the training can easily become stuck in certain local optimum situations, resulting in wrong estimations with averaged or smoothed values. To avoid the local optimum and obtain effective predictions for different samples, we design a standard deviation-based loss term (ℒ_*std*_) as a regularization method. Specifically, ℒ_*std*_ requires the standard deviation of the prediction results in a batch to approach that of the regression ground truth values in this batch, which can be expressed as follows:

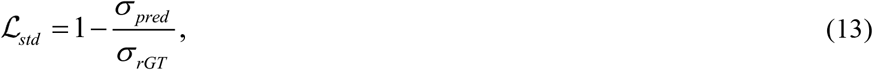

where *σ*_*pred*_ and *σ*_*rGT*_ indicate the standard deviations of the predicted values and regression ground truth values in the single batch, respectively. Then, the total loss function is:

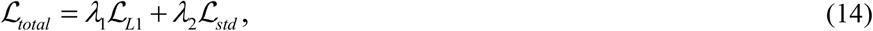

where *λ*_1_ and *λ*_2_ are two coefficients for weighting different terms for stable and efficient convergence.

### 2.5 Implementation and evaluation

#### The selection of coefficients for MIhd

There are three coefficients (i.e., *ε, δ*, and *ω*) to be confirmed when applying MIhd. These hyper-parameters may expect different values for different objects due to different human behaviors for different objects. For example, the influence of T1 errors (corresponding to *ε*) and boundary distance (corresponding to *ω*) may vary according to the distribution of errors. In addition, intuitively speaking, human readers tend to accept relatively larger *δ* for large objects (e.g., lung) than for small objects (e.g., esophagus) in 2D images.

Therefore, these three coefficients require specific optimization for each object. Specifically, we decide *δ* for each object by comparing two sets of manual segmentation masks for the same CT images from our previous study [33]. The HD values between these two sets were calculated, resulting in the average HD for each object as the value of *δ*. In contrast, for the coefficients *ε* and *ω*, hyper-parameter grid searching was conducted for each organ such that the ultimately decided *ε* and *ω* could maximize the correlation between MIhd and *nTm*.

#### Implementation of deep learning-based models

The three deep learning-based pipelines were implemented using the PyTorch framework in the Ubuntu system. All the models can be individually achieved using a single Nvidia RTX A5000 GPU with a batch size of 8 and a learning rate of 1*e* − 4. Each training lasts for 200 epochs. The coefficients in the total loss function are 10.0 and 0.5 for *λ*_1_ and *λ*_2_, respectively.

#### Dataset splitting

For all of the following correlation analysis experiments, all of the collected samples for each object are used to calculate the correlation between each explicitly defined segmentation metric with the mending effort representative *nTm*.

For all of the following regression analysis experiments, including both classical regression and deep learning-based regression, five-fold cross-validation is applied and the mean performance from the five-fold splitting is calculated. The five-fold splitting for each object remains the same for all experiments using different segmentation metrics.

## 3. Results

This study was conducted based on the data obtained from a previous multi-center prospective study [33], following approval from the Institutional Review Board (IRB) at the Hospital of the University of Pennsylvania, the single IRB of record, along with a Health Insurance Portability and Accountability Act waiver.

### 3.1 Explicit models

#### Correlation analysis

To study how well each explicit segmentation metric could represent the effort required for mending auto-segmentations, correlation analysis was conducted. Specifically, for each object and each of the compared metrics (i.e., mDC, sHD, surDC, sAPL, or MIhd), the correlation between the metric value with the *nTm* records from different institutions was calculated. The Pearson correlation coefficient was applied for both positive correlation and negative correlation, which is presented in Table 3 and Table 4. Based on the definitions of the above metrics, mDC, surDC, and MIhd are expected to have negative correlations with *nTm*. In contrast, sHD and sAPL are expected to have positive correlations with *nTm*.

**Table 3.** Correlation coefficients between each segmentation metric with the mending effort *nTm* for sparse objects. The highest and the second highest correlation coefficients for each object from each institution are bold and underscored, respectively.

| Object<br>Metric | LSmG |  |  |  |  | Mnd |  |  |  |  | OHPH |  |  |  |  |
| --- | --- | --- | --- | --- | --- | --- | --- | --- | --- | --- | --- | --- | --- | --- | --- |
|  | mDC | sHD | surDC | sAPL | MIhd | mDC | sHD | surDC | sAPL | MIhd | mDC | sHD | surDC | sAPL | MIhd |
| Institution A | -0.02 | 0.18 | -0.12 | **0.67** | <u>-0.34</u> | 0.19 | 0.10 | -0.09 | <u>0.27</u> | **-0.48** | -0.37 | <u>0.38</u> | -0.18 | 0.08 | **-0.67** |
| Institution B | -0.14 | 0.24 | **-0.54** | <u>0.47</u> | -0.34 | -0.09 | <u>0.28</u> | 0.03 | -0.04 | **-0.51** | -0.33 | <u>0.50</u> | -0.49 | 0.38 | **-0.56** |
| Institution C | -0.41 | 0.36 | -0.53 | **0.61** | <u>-0.55</u> | <u>-0.27</u> | 0.15 | 0.05 | -0.11 | **-0.35** | -0.19 | 0.18 | <u>-0.40</u> | **0.59** | -0.31 |
| All institutions | -0.32 | 0.32 | -0.45 | **0.55** | <u>-0.44</u> | <u>-0.28</u> | 0.19 | -0.13 | 0.10 | **-0.43** | -0.23 | <u>0.26</u> | -0.18 | 0.17 | **-0.46** |
| Object<br>Metric | CtTr |  |  |  |  | CtSC |  |  |  |  |  |  |  |  |  |
|  | mDC | sHD | surDC | sAPL | MIhd | mDC | sHD | surDC | sAPL | MIhd |  |  |  |  |  |
| Institution A | 0.20 | **0.70** | -0.25 | <u>0.62</u> | -0.59 | -0.00 | <u>0.48</u> | -0.22 | 0.34 | **-0.58** |  |  |  |  |  |
| Institution B | **-0.14** | 0.09 | 0.05 | -0.11 | <u>-0.10</u> | 0.12 | -0.36 | 0.22 | -0.45 | 0.35 |  |  |  |  |  |
| Institution C | -0.04 | 0.19 | <u>0.22</u> | 0.05 | **-0.34** | **-0.72** | 0.20 | <u>-0.39</u> | 0.12 | -0.19 |  |  |  |  |  |
| All institutions | 0.12 | **0.44** | -0.11 | <u>0.40</u> | -0.37 | -0.04 | <u>0.10</u> | -0.02 | 0.08 | **-0.21** |  |  |  |  |  |

**Table 4.** Correlation coefficients between each segmentation metric with the mending effort *nTm* for non-sparse objects. The highest and the second highest correlation coefficients for each object from each institution are bold and underscored, respectively.

| Object<br>Metric | RLg |  |  |  |  | Hrt |  |  |  |  |
| --- | --- | --- | --- | --- | --- | --- | --- | --- | --- | --- |
|  | mDC | sHD | surDC | sAPL | MIhd | mDC | sHD | surDC | sAPL | MIhd |
| Institution A | -0.36 | **0.61** | -0.47 | 0.41 | <u>-0.53</u> | **-0.50** | 0.33 | -0.35 | 0.34 | <u>-0.50</u> |
| Institution B | 0.03 | **0.21** | 0.05 | -0.08 | <u>-0.17</u> | -0.25 | <u>0.44</u> | -0.41 | 0.32 | **-0.56** |
| Institution C | -0.28 | 0.44 | **-0.62** | 0.43 | <u>-0.46</u> | -0.43 | **0.54** | -0.24 | 0.01 | <u>-0.44</u> |
| All institutions | -0.27 | **0.53** | -0.44 | 0.44 | <u>-0.46</u> | -0.42 | **0.47** | -0.27 | 0.18 | <u>-0.46</u> |

The correlation analysis for sparse objects is presented in Table 3. The results appear to vary for different institutions. For example, when correcting auto-segmentations of CtSC, the reader’s behavior from Institution C appears most related to area-based metrics (e.g., mDC), while readers from other institutions may depend more on boundary distance and boundary length for correction. The results also seem to differ for different objects as well. For example, by observing the results related to Institution B, all the metrics do not show very strong correlation with *nTm* when correcting CtTr segmentations, but some higher correlation coefficients are found when mending other objects.

Generally speaking, MIhd may be a good choice to indicate the mending effort for sparse objects. By considering the highest and the second highest correlation coefficients as the strong correlation coefficients, MIhd shows strong correlation for 4 objects, 3 objects, 3 objects, and 3 objects based on samples from Institution A, Institution B, Institution C, and All Institutions, respectively. In contrast, the quantities of strong correlation coefficients obtained by sAPL are 3, 1, 2, and 2, the quantities of strong correlation coefficients obtained by sHD are 3, 2, 1, 3, and mDC and surDC obviously perform worse.

Similarly, the correlation analysis for non-sparse objects is shown in Table 4. The correlation values of segmentation metrics still vary for different institutions and different objects. For example, compared with readers in other institutions, mending effort from readers from Institution B may be less related to area-based errors. Even in the same institution, for example, in Institution A, human readers’ behaviors show different patterns in terms of boundary distance-based errors when correcting RLg and Hrt auto-segmentations.

When we choose the highest and the second highest correlation coefficients for each institution for each object as the strong correlation coefficients, we can summarize that MIhd and sHD are two appropriate metrics for estimating the auto-segmentation mending effort for non-sparse objects. As shown in Table 4, MIhd and sHD generally outperform other metrics for both RLg and Hrt.

#### Classical regression analysis

As mentioned before, the regression analysis was conducted to directly verify the capacity of mending effort prediction using analytic metrics. Mean absolute error (MAE) was applied to measure the difference between the predicted value and the manually recorded *nTm* (i.e., ground truth for the regression experiment). Because *nTm* is a normalized value as shown in Eq. (1) indicating the relative required mending effort or relative mending difficulty for each human reader, the MAE value in the regression analysis can be expressed as a percentage to evaluate the regression performance intuitively. As such, the mean value and the standard deviation results of the five-fold cross-validation are presented in Table 5 and Table 6.

**Table 5.**
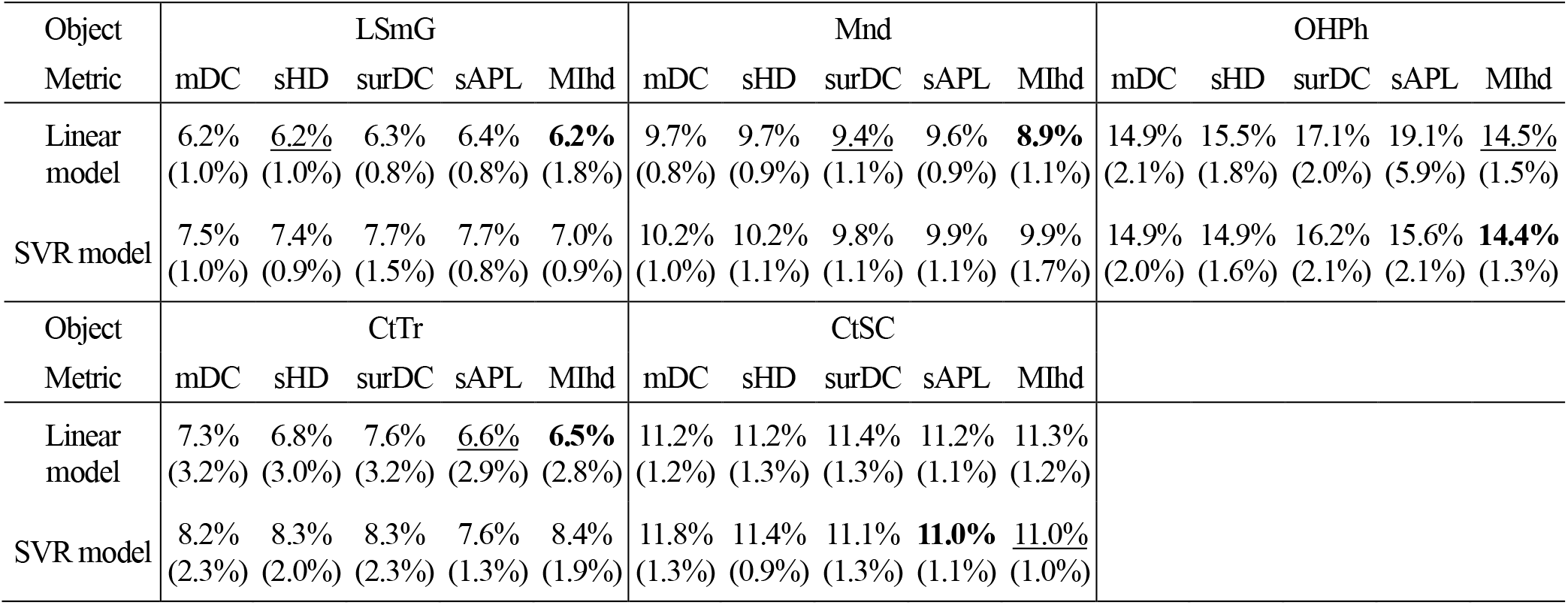
Classical regression performance for sparse objects. MAEs are presented as percentages. The standard deviations of the five-fold cross-validation are provided in brackets. The smallest and the second smallest errors for each object are bold and underscored, respectively.

| Object<br>Metric | LSmG |  |  |  |  | Mnd |  |  |  |  | OHPh |  |  |  |  |
| --- | --- | --- | --- | --- | --- | --- | --- | --- | --- | --- | --- | --- | --- | --- | --- |
|  | mDC | sHD | surDC | sAPL | MIhd | mDC | sHD | surDC | sAPL | MIhd | mDC | sHD | surDC | sAPL | MIhd |
| Linear model | 6.2%<br>(1.0%) | <u>6.2%</u><br>(1.0%) | 6.3%<br>(0.8%) | 6.4%<br>(0.8%) | **6.2%**<br>(1.8%) | 9.7%<br>(0.8%) | 9.7%<br>(0.9%) | <u>9.4%</u><br>(1.1%) | 9.6%<br>(0.9%) | **8.9%**<br>(1.1%) | 14.9%<br>(2.1%) | 15.5%<br>(1.8%) | 17.1%<br>(2.0%) | 19.1%<br>(5.9%) | <u>14.5%</u><br>(1.5%) |
| SVR model | 7.5%<br>(1.0%) | 7.4%<br>(0.9%) | 7.7%<br>(1.5%) | 7.7%<br>(0.8%) | 7.0%<br>(0.9%) | 10.2%<br>(1.0%) | 10.2%<br>(1.1%) | 9.8%<br>(1.1%) | 9.9%<br>(1.1%) | 9.9%<br>(1.7%) | 14.9%<br>(2.0%) | 14.9%<br>(1.6%) | 16.2%<br>(2.1%) | 15.6%<br>(2.1%) | **14.4%**<br>(1.3%) |
| Object<br>Metric | CtTr |  |  |  |  | CtSC |  |  |  |  |  |  |  |  |  |
|  | mDC | sHD | surDC | sAPL | MIhd | mDC | sHD | surDC | sAPL | MIhd |  |  |  |  |  |
| Linear model | 7.3%<br>(3.2%) | 6.8%<br>(3.0%) | 7.6%<br>(3.2%) | <u>6.6%</u><br>(2.9%) | **6.5%**<br>(2.8%) | 11.2%<br>(1.2%) | 11.2%<br>(1.3%) | 11.4%<br>(1.3%) | 11.2%<br>(1.1%) | 11.3%<br>(1.2%) |  |  |  |  |  |
| SVR model | 8.2%<br>(2.3%) | 8.3%<br>(2.0%) | 8.3%<br>(2.3%) | 7.6%<br>(1.3%) | 8.4%<br>(1.9%) | 11.8%<br>(1.3%) | 11.4%<br>(0.9%) | 11.1%<br>(1.3%) | **11.0%**<br>(1.1%) | <u>11.0%</u><br>(1.0%) |  |  |  |  |  |

**Table 6.** Classical regression performance for non-sparse objects. MAEs are presented as percentages. The standard deviations of the five-fold cross-validation are provided in brackets. The smallest and the second smallest errors for each object are bold and underscored, respectively.

| Object<br>Metric | RLg |  |  |  |  | Hrt |  |  |  |  |
| --- | --- | --- | --- | --- | --- | --- | --- | --- | --- | --- |
|  | mDC | sHD | surDC | sAPL | MIhd | mDC | sHD | surDC | sAPL | MIhd |
| Linear model | 10.7%<br>(3.0%) | <u>9.6%</u><br>(2.1%) | 10.4%<br>(2.7%) | 10.6%<br>(2.5%) | 10.9%<br>(2.1%) | 14.7%<br>(3.7%) | 14.0%<br>(3.0%) | 14.2%<br>(3.0%) | 13.6%<br>(2.3%) | **13.4%**<br>(2.3%) |
| SVR model | 10.8%<br>(3.6%) | **9.5%**<br>(2.2%) | 10.4%<br>(3.0%) | 10.7%<br>(2.7%) | 10.3%<br>(1.5%) | 15.3%<br>(2.8%) | 13.9%<br>(2.8%) | 14.1%<br>(3.0%) | <u>13.4%</u><br>(2.0%) | 14.6%<br>(2.3%) |

The classical regression results for sparse objects are found in Table 5. The two smallest errors in terms of each object are additionally emphasized using bold and underscore. According to Table 5, the linear regression model may be slightly better than the SVR model. Although the SVR model outperforms the linear model for OHPh and CtSC, the difference is trivial. In contrast, the gaps for the other three objects are obvious.

Also, the estimation performance appears to vary a lot for different objects. For example, when estimating the effort for mending LSmG’s auto-segmentation results, all of the regression models based on either metric obtain an average MAE of <8%. However, all of the average MAE values are larger than 14% for the object OHPh.

Moreover, by comparing different auto-segmentation metrics, one can conclude that MIhd can be used to generally better estimate *nTm* compared to other metrics for sparse objects. For all of the five objects in Table 5, the relatively small regression errors (i.e., emphasized values) can be obtained by taking MIhd as the input.

The classical mending effort regression experiment results for non-sparse objects are shown in Table 6. From the two representative objects, we easily observe different performances between different objects. In addition, it seems that sHD and MIhd can lead to the best regression performance using the appropriate model for RLg and Hrt, respectively.

The above correlation analysis and classical regression analysis are consistent to some extent. Regarding the answer to the question regarding which auto-segmentation metric is best for indicating the mending effort, it appears to differ for sparse and non-sparse objects. In both types of experiments, MIhd generally outperforms the other four metrics when targeting sparse objects. In contrast, for non-sparse objects, it is not easy to decide the best choice between MIhd and sHD. Both of them may be acceptable.

### 3.2 Implicit models

As mentioned before, in addition to the five explicitly defined auto-segmentation analytic metrics, we also try to verify the feasibility of deep learning-based mending effort estimation (i.e., implicit models), which avoids the strict error classification and may implicitly fit human behaviors.

Three types of models have been introduced before, corresponding to different types of inputs. The same five-fold cross-validation as in the classical regression experiment was conducted, which is evaluated using MAE in percentage as well. The regression performance results are found in Table 7. As the baseline, the smallest MAE obtained in the classical regression experiment for each object is also displayed in Table 7.

**Table 7.** Deep learning-based regression performance. MAEs are presented as percentages. The standard deviation results of the five-fold cross-validation are provided in brackets. The smallest and the second smallest errors for each object are bold and underscored, respectively.

| Object | LSmG | Mnd | OHPH | CtTr | CtSC | RLg | Hrt |
| --- | --- | --- | --- | --- | --- | --- | --- |
| Best classical regression | 6.2%<br>(1.8%) | 8.9%<br>(1.1%) | 14.4%<br>(1.5%) | 6.5%<br>(2.8%) | 11.0%<br>(1.1%) | 9.5%<br>(2.2%) | 13.4%<br>(2.0%) |
| Auto+GT | <u>3.9%</u><br>(1.1%) | **6.8%**<br>(1.4%) | **12.7%**<br>(2.6%) | 6.4%<br>(1.5%) | 9.4%<br>(2.4%) | <u>9.1%</u><br>(2.6%) | **12.9%**<br>(0.6%) |
| Auto+GT+CT | **2.9%**<br>(0.9%) | 8.0%<br>(2.6%) | <u>13.2%</u><br>(2.8%) | <u>6.2%</u><br>(0.8%) | **8.9%**<br>(1.4%) | **8.7%**<br>(2.0%) | 13.0%<br>(1.8%) |
| Auto+CT | 4.0%<br>(1.8%) | <u>7.6%</u><br>(2.6%) | 13.3%<br>(2.7%) | **5.6%**<br>(1.5%) | <u>9.2%</u><br>(2.2%) | 9.2%<br>(2.0%) | <u>13.0%</u><br>(2.5%) |

Table 7 shows the performance of estimating the mending effort for both sparse and non-sparse objects’ auto-segmentations using deep learning models. One can observe that all of the deep learning models surpass the best classical regression results for all seven objects in terms of the average MAE. For example, for the object of LSmG, the average MAE could be greatly reduced from 6.2% to 2.9% by using the “Auto+GT+CT” pipeline. The experiments demonstrate that it is feasible to replace explicit models by implicit deep learning-based models to evaluate the auto-segmentations from the viewpoint of clinical value.

Meanwhile, even with deep learning-based models, different estimation performances can still be observed for different objects. For example, the estimation for LSmG (with average MAE of 2.9%-4.0%) is more accurate than that for OHPh (with the average MAE of 12.7%-13.3%). Such variances among objects are closely related to the complexity of objects, variant clinical standards among different institutions, different subjective experiences, and different correction tools, which influence *nTm* labeling and also model training. Although the regression methods and theories are totally different, objects with relatively high classical regression errors tend to show relatively high deep learning-based regression errors, indicating that the differences among objects may be more related to the data collection instead of the regression algorithms.

In addition, by comparing different deep learning-based models, “Auto+GT” model and “Auto+GT+CT” model generally perform similarly. Each of the models obtains the smallest average MAE values for three objects and the second smallest average MAE values for two objects. Specifically, “Auto+GT” model achieved the best estimation for Mnd, OHPh, and Hrt, while showing slightly poorer results compared to the “Auto+GT+CT” model for LSmG, CtSC, and RLg. Although “Auto+GT+CT” model utilizes the most input information, it may become more difficult to achieve effective feature extraction for all cases. Therefore, it is reasonable that the “Auto+GT+CT” cannot always outperform “Auto+GT” model as observed in our experiments.

It is important to notice the performance of “Auto+CT” model. Although the average MAE values indicate that “Auto+CT” model generally performs slightly worse than the other two counterparts except for CtTr, there are often no significant differences. For Mnd, CtTr, CtSC, and Hrt, the “Auto+CT” model can even obtain the smallest or the second smallest errors. Although the classical regression methods were implemented with the utilization of GT segmentations for segmentation metric calculation, the “Auto+CT” model could be more accurate than all the classical regression models for all objects. As the “Auto+CT” pipeline avoids the requirement of manually labeled ground truth segmentation, this method provides the possibility to evaluate auto-segmentations with fewer labels but with comparable clinical value, potentially enabling its application in many clinical scenarios. For example, the demands for manual GT segmentations could be reduced during developing or evaluating auto-segmentation algorithms, resulting in lower cost and higher efficiency.

## 4. Discussion

### 4.1 The normalization of *Tm*

As mentioned in the Method Section, the mending effort Tm was normalized according to each reader’s records to represent the relative mending effort individually, which is a practical way to alleviate the influence from varying personal experience and center-specific situations. In our study, compared with directly applying *Tm*, the utilization of *nTm* could generally improve the correlation and prediction performance. For example, according to Table 3 and Table 4, the correlation coefficients between *nTm* and sHD are 0.44, 0.53, and 0.47, for CtTr, RLg, and Hrt, respectively. However, when directly applying Tm to indicate the mending effort, these values decrease to 0.40, 0.52, and 0.41. Similar degradations may generally obtained for different objects using various metrics. Therefore, it is useful to adopt *nTm* for the purpose of mending effort evaluation and estimation.

### 4.2 Contouring behavioral differences among different institutions

Even with the normalization of *Tm*, different institutions may experience fluctuations of the values as seen in Tables 3-6, which can be attributed to two types of issues. Firstly, the center-specific contouring culture, including the personal experiences, correction software, and subjective acceptability levels, can directly lead to different mending times. Second, there are no standardized body region definitions for some complex objects, such as CtSC, used in clinical practice. Thus, institutional clinical standards vary for these objects. The differences among institutions could thereby influence the experiment performance when mixing the multi-source data, for both explicit models and implicit models. With the use of standardized definitions in the future for mending contours and recording mending time, the approaches proposed in this study should yield much more accurate prediction results.

Some examples from OHPh and Hrt samples are presented below for demonstrating the different clinical standards used at different institutions. Specifically, all the GT segmentations were labeled by our group following the unique standard based in large part on published consensus guidelines. In contrast, we further invited readers from the other three institutions to fully manually segment the OHPh and Hrt objects following their own clinical standards. Therefore, we are able to compare if their clinical standards are consistent with our GT.

We firstly compare the two types of segmentations, i.e., the ground truth segmentation with the unique standard and the clinical segmentation from three institutions, using both 2D and 3D Dice coefficient metrics. As shown in Table 8, for the clinical segmentation from each institution, the minimal, maximal, and mean metric values followed by the standard deviation (std) values are presented.

**Table 8.** Dice coefficient values from comparing the clinical contours from three institutions with the unique ground truth contour.

| Object | Metric | 2D Dice |  |  | 3D Dice |  |  |
| --- | --- | --- | --- | --- | --- | --- | --- |
|  | Institution | Min value | Max value | Mean (std) value | Min value | Max value | Mean (std) value |
| Hrt | Institution A | ~0 | 0.99 | 0.93 (0.09) | 0.88 | 0.96 | 0.93 (0.02) |
|  | Institution B | ~0 | 0.99 | 0.89 (0.11) | 0.78 | 0.96 | 0.89 (0.04) |
|  | Institution C | ~0 | 0.99 | 0.92 (0.11) | 0.84 | 0.94 | 0.90 (0.02) |
| OHPH | Institution A | ~0 | 0.91 | 0.49 (0.21) | 0.25 | 0.56 | 0.39 (0.08) |
|  | Institution B | 0.14 | 0.92 | 0.67 (0.13) | 0.48 | 0.72 | 0.61 (0.07) |
|  | Institution C | 0.14 | 0.89 | 0.65 (0.13) | 0.49 | 0.71 | 0.57 (0.07) |

According to Table 8, we observe that different institutions indeed show different consistency with our ground truth. For Hrt, clinical segmentations from Institution B obtain the lowest mean 2D Dice and mean 3D Dice values with the largest standard deviations, indicating that the contour definition of Hrt at Institution B may be different from that of the other two institutions and our ground truth. This inconsistency phenomenon may be more significant for OHPh. For example, compared with our ground truth, cases from Institution B and Institution C could result in the minimal 3D Dice of around 0.5, while the most different case in Institution A obtains the value of 0.25, which reduces almost half. With a much different lower limit (i.e., minimal 3D Dice), upper limit (i.e., maximal 3D Dice), and average performance (i.e., mean 3D Dice), readers from Institution A appear to have a very different standard to delineate OHPh compared to readers from Institutions B and C.

The differences between each institution and our ground truth can also be qualitatively observed in Fig. 7. Again, selecting Hrt and OHPh as representative objects, clinical segmentations from three institutions are compared with the ground truth segmentation with different colors for demonstrating different regions. Compared with the ground truth segmentation, some examples showing relatively nice agreement and relatively poor agreement are presented.

**Fig. 7.**
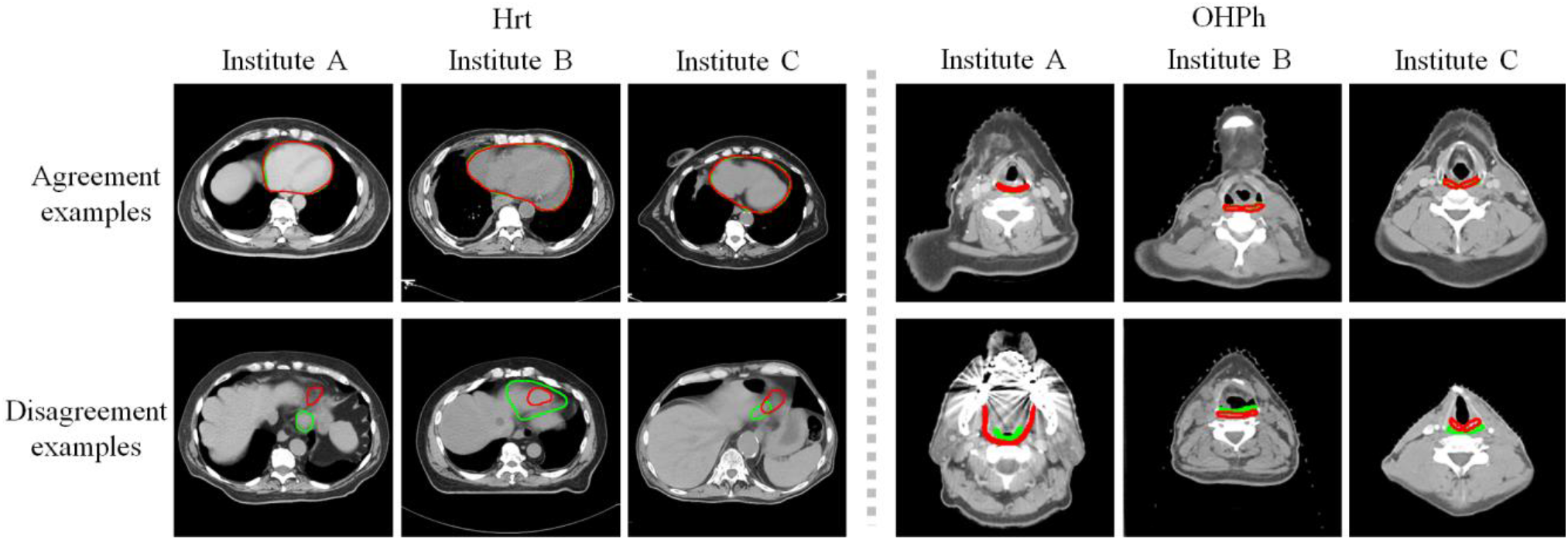
Representative examples showing the relative agreement and relative disagreement between different institutions’ clinical segmentation with the GT for objects Hrt and OHPh. Red and green indicate the clinical contour and the GT contour, respectively.

According to Fig. 7, Hrt labeling examples from Institution B show different error patterns from those of Institutions A and C, as the representative example shows more FN errors. Moreover, it is evident that readers from Institution A tend to label a larger OHPh region than readers from the other two institutions and our group, resulting in the poor values in Table 8.

Table 8 and Fig. 7 have demonstrated some contouring behavioral differences among different institutions. This is a very practical and valuable issue to highlight in both this study with multi-center prospective data and in the real-world clinical routine. Although we have normalized the recorded *Tm* to alleviate the influence of *Tm* recording, the different standards of object delineation or correction can still impact the metric calculation and deep learning model training in our study.

### 4.3 Discussion about explicit models

In this study, we compared five well-defined auto-segmentation metrics, including the existing mDC, sHD, surDC, sAPL, and the newly proposed MIhd. These metrics explicitly classify and count different types of errors, showing different performances to estimate the effort required for manually mending the auto-segmentations.

From the perspective of design motivation, the proposed MIhd has several specific considerations. Firstly, it adopts the segmentation boundary length-based concepts. Since human readers often correct auto-segmentations based on boundary length on 2D slices, it is reasonable to simulate the human behaviors for better indicating the clinical value. Second, MIhd considers the influence of T1 errors, discusses two types of other more serious errors, and introduces a fuzzy version, which are usually ignored in other boundary length-based metrics. The removal efficiency for these isolated T1 errors is usually related to the annotation software. In addition, MIhd also combines a variant of HD, the boundary distance-based concept, and the object size. On the one hand, as shown in the correlation analysis and classical regression analysis, the relevance between sHD and *nTm* is generally not weak. On the other hand, it is reasonable that a larger object with larger intersecting surface and more slices tends to need more time for auto-segmentation correction (e.g., the 3D sizes of the esophagus vary a lot among patients).

However, according to the correlation analysis and the classical regression analysis, it is still difficult to answer the question regarding which auto-segmentation metric can always robustly indicate the mending effort for all objects. The performance fluctuations among objects and institutions may be heavily related to the above-mentioned institution standards issue. Although we cannot conclude an absolutely consistent choice, since MIhd generally outperforms other metrics for sparse objects and shows comparable performance with sHD for non-sparse objects, MIhd may generally be a good choice for estimating the required mending effort.

### 4.4 Discussion about deep learning-based implicit estimation

Human behaviors are extremely complicated to be strictly summarized in a simple analytical formula, even for MIhd with several different considerations. As such, for estimating the mending effort, the above explicitly defined auto-segmentation metrics show some limitations in simulating the manual correction procedure. Although without explicit recognition of errors and human behaviors, deep learning architectures with great fitting capacity are expected to better perceive the auto-segmentations and the corrections.

The results of the deep learning-based regression experiments in Table 7 verify our hypothesis that the deep learning-based method can potentially work as an implicit metric for indicating the mending effort. The estimation performance of deep learning-based pipelines can outperform classical regression models based on explicit metrics for all of the studied objects. Therefore, these deep learning-based pipelines have the potential to replace explicit metrics for evaluating auto-segmentations and even for guiding the development of auto-segmentation algorithms, thereby boosting those auto-segmentation techniques that have greater clinical value.

Moreover, the “Auto+CT” model is unique among the three mentioned deep learning-based models and the explicitly defined metrics, since the “Auto+CT” model does not require the manual annotation of the ground truth segmentation and yet obtains even better estimation performance than the explicitly defined metrics. One the one hand, the avoidance of GT segmentation could tremendously reduce the workload necessary from the clinical side when developing auto-segmentation methods for specific tasks. Since the “Auto+CT” model does need the GT segmentation when evaluating auto-segmentations, the prediction could possibly work as a loss function for an unsupervised learning scheme, benefitting the utilization of numerous unlabeled medical data. On the other hand, the model could be introduced into the clinical routine to automatically prompt the quality of auto-segmentations without manual GT, helping readers to correct the auto-segmentations more efficiently and effectively.

### 4.5 Challenges and future work

Some challenges and limitations exist for this study using multi-center data. Firstly, in terms of the data collection from different institutions, there might be the following challenges. The variation of institutional contouring standards is a practical challenge that could impact the regression model training. Therefore, creating a universal standard through standardized definitions is important for this proposed study and also for many other clinical applications. Also, the mending effort *Tm* is recorded in terms of the whole 3D object in this study. Since the manual correction is usually based on 2D slices, more precise mending time records for all 2D slices might be more appropriate to analyze human behaviors, which requires much more complicated data collection procedures that can be conducted in the future. Furthermore, the limited dataset with *Tm* could surely limit the robustness and generalization of regression models and conclusions. It is extremely challenging to collect such information from experts and multiple centers with a reasonable cost.

Second, although MIhd generally performs well in indicating the mending effort in this study, there are some limitations and some further expectations. Due to the design of tolerant boundary length-based error counting and the element definition of pixel edges, the calculation of MI (or MIhd) is relatively slow, hindering the potential of using MI as a loss function. In addition, MI is an object-specified metric with specifically grid-searched parameters. In other words, the high mending effort estimation performance of MIhd is partly at the cost of accessibility. Therefore, further improvements to the metric design are still expected to improve its accessibility and applicability.

Lastly, the deep learning-based experiments mainly focused on validating the feasibility of applying neural networks to estimate the clinical mending effort for auto-segmentations and the potential of the “Auto+CT” pipeline. Therefore, this study does not heavily explore all of the possible network architectures or attempt to propose novel network blocks. In other words, more algorithm-based innovations could be explored in the future to optimize the estimation performance and to benefit downstream applications.

## 5. Conclusion

This paper discusses the abilities of different segmentation metrics for forecasting the effort required for manually mending auto-segmentations on medical images. Instead of defining certain similarities between the auto-segmentation with the GT segmentation when designing metrics, we pay more attention to the clinical value of the auto-segmentation. Therefore, datasets generated in a previous multi-center study [33] were utilized in this paper, consisting of auto-segmentations, ground truth segmentations (additionally labeled by our group with unique definitional standards), and the time needed for manually correcting each auto-segmented object. Explicit models with correlation analysis and classical regression analysis, and implicit models (i.e., deep learning-based regression analysis) were conducted.

In addition to the area-based metric mDC, the boundary distance-based metric sHD, and the boundary length-based metrics surDC and sAPL, a novel segmentation metric MI as well as its variant MIhd were proposed with the consideration of the boundary length-based concept, the boundary-distance concept, and the influence of object size. The correlation analysis and classical regression analysis indicate that sHD might be a good choice for non-sparse objects, while MIhd could be a generally excellent metric for all objects to indicate the clinical value of auto-segmentations.

Three types of deep learning-based models can work as implicit means to evaluate auto-segmentation results, showing even better performance to estimate the auto-segmentation mending effort than those well-defined metrics (i.e., mDC, sHD, surDC, sAPL, and MIhd). It is feasible to use neural networks to evaluate auto-segmentation results or even guide the training. Moreover, it is possible to avoid manually labeled ground truth segmentation when evaluating auto-segmentations using the “Auto+CT” model from the perspective of clinical demands (i.e., mending effort). We hope that this work will inspire the development of auto-segmentation techniques with more practical concerns for clinical usability.

## Data Availability

All data produced in the present study are currently not publicly available.

## Footnotes

1 Notably, per-study segmentation cost in P3 is what matters usually so long as the training cost is not prohibitive and impractical, and thus P3 reduces essentially to computational cost per study to perform auto-segmentation.

